# Does prior healthcare experience predict success on clinical courses and add value to admissions processes?

**DOI:** 10.1101/2021.07.06.21258905

**Authors:** Claire Darling-Pomranz, James Gray, David Spencer

## Abstract

**Objectives:** This work sought to assess whether prior clinical experience provided any guide to likely course achievement from three completed cohorts of Physician Associates at the University of Sheffield.

**Methods:** Sixty students who entered the PA course at TUoS since it began in 2016 were included in the study. Each students’ original course application was reviewed for healthcare experience and mapped against first sit examination scores. Statistical analysis was undertaken with a two-tailed t-test.

**Results:** No correlation was found between previous healthcare experience and performance in examinations. Students with previous healthcare experience performed slightly worse than those without in the OSCE examination but not at a level of statistical significance.

**Conclusions:** The use of clinical experience as part of the criteria of entry does not predict success on a Physician Associate course. We support the position of the 2010 Ottawa conference that quality assured methodologies along with objective cut offs for previous academic attainment are the most appropriate way to select students for clinical courses.

## Introduction

The selection of students for clinical courses has changed over the years. Whilst most courses retain an interview these may be in different formats and, in the UK, many use aptitude testing such as the UKCAT (United Kingdom Clinical Aptitude Test).

Physician Associate (PA) courses in the UK are almost exclusively postgraduate taught courses taking graduates with a life science degree. Courses have variable entry requirements for their courses based upon previous degree classifications and, in some cases, prior clinical experience. A statistically significant correlation between prior clinical experience and course performance could provide an important subcomponent to admission criteria.

An initial scoping literature search found no papers which had looked at this area. Ethics approval was granted through the University of Sheffield (reference 037170).

## Methods

All students who entered the PA course at TUoS between 2016 and 2018 were included totalling sixty students. Each students’ course application was reviewed to determine whether they were employed within the healthcare sector in a patient facing or non-patient facing role. Shadowing, volunteering and previous placement experience were not considered significant healthcare experience.

The PA programme has three primary summative assessments during the two year course. Single Best Answer (SBA) examinations at the end of years one and two and an Objective Structured Clinical Examination (OSCE) at the end of year two. The results for each student for the first sit of each of these exams was recorded. The examinations utilise the Angoff method to determine the pass mark^2^ leading to different pass marks for each assessment.

To allow comparison, all exam scores were rescaled to a pass mark of 50% using a linear rescaling method.

In order to test the assumption of normal distribution of results we carried out a Kolmogorov-Smirnov goodness of fit test which confirmed this. F-tests showed no evidence of a difference in variances between groups.

## Results

TUoS had twenty students accepted to the PA course per cohort. There were a number of students who failed either the year one SBA or the first attempt at the OSCE. Sixty year one SBA and fifty three year two SBA and OSCE results were included.

Mean scores for each assessment were calculated separately for those with no healthcare experience and those with experience of either sort (patient facing or non-patient facing) together with the minimum and maximum scores for each assessment.

**Table 1:** Comparison of those students with healthcare experience and those with none

|  | **Previous healthcare experience** |  |
| --- | --- | --- |
|  | **No** | **Yes** |
| **# of students** | **28** | **32** |
| Mean score in year 1 SBA | 63.54 | 62.87 |
| Maximum score in year 1 SBA | 81.06 | 82.00 |
| Minimum score in year 1 SBA | 34.95 | 40.72 |
| **# of students** | **24** | **29** |
| Mean score in year 2 SBA | 67.99 | 65.25 |
| Maximum score in year 2 SBA | 81.65 | 81.32 |
| Minimum score in year 2 SBA | 55.14 | 50.97 |
| Mean score in OSCE | 69.53 | 65.43 |
| Maximum score in OSCE | 81.05 | 83.03 |
| Minimum score in OSCE | 51.61 | 46.68 |

A two-tailed t-test for each exam revealed no significant differences between the mean scores of the two groups.

Some of the students failing the first sitting of the year one SBA exam did not progress into year two and their lower scores could have affected the analysis. We re-did the year one SBA results analysis only including those students who passed the exam with no change in the findings.

It was noted that there was almost a significant difference (p = 0.057) between the two groups OSCE examination scores (mean score for those without experience 69.53%, mean for those with experience 65.43%) suggesting prior experience led to worse OSCE performance. We further analysed this by splitting the healthcare experience category into patient facing and non-patient facing experience with a similar difference remaining between those with patient facing experience and those with no experience (p = 0.067). Three students failed the OSCE at first attempt so we carried out additional analysis removing their results. The difference in scores remained and was close to statistical significance (p = 0.072). This phenomenon was further analysed by considering each year cohort separately. Although this was with relatively small numbers it demonstrated the same pattern as a year on year trend.

## Discussion

A 1998 British Medical Journal article noted that clinical experience on a medical course was not itself a predictor of performance in final examinations but that wider experience did relate to more strategic and deep learning styles than those with less experience^3^. Work in 2003 looked at non-interview or testing measures in the selection process including personal statements and letters of recommendation with no evidence to show that they added anything to an appropriate interview process^4^. Authors looking at psychology trainees found that it was A-level and degree performance which acted as key predictors of success^5^. The Ottawa conference in 2010 produced a consensus statement and recommendations for selection focused on following quality assurance processes already expected within course assessment^6^. They noted the value of aptitude testing, and multiple mini interviews with a core evidence base as part of appropriate measurement. United Kingdom studies of the UKCAT have shown a lack of consensus as to whether scores predict success with one showing a correlation^7^ whilst another showed none^8^. Previous work looking at both the cognitive and non-cognitive elements of UKCAT and their predictive value in success showed individual predictive value of any one element to be weak^9^ though noted previous work that showed that even weak correlations can add value as predictors of success where the application to selection ratio is large^10^.

Our results suggested that students with prior patient facing healthcare experience did slightly worse in OSCE exams than their peers. It is unclear whether this is related to developed habits or previously developed heuristics but is an area for potential future research.

This study has a number of limitations. All of the subjects were from a single university and represent relatively small numbers for statistical analysis. In addition whilst all students had at least a 2:1 in their primary degree, providing a degree of academic homogeneity, the degrees themselves would have had a variety of content which may have affected the results.

## Conclusion

Our results suggest that use of clinical experience as part of the criteria of entry does not predict success on a Physician Associate course. We support the position of the Ottawa conference that quality assured methodologies along with objective cut offs for previous academic attainment in appropriate subjects are the most appropriate way to manage the selection of students for the vocational clinical courses.

## Data Availability

The data for this trial is secure due to sensitivity of personally identifiable information. Anonymised data could be provided on request

